# Assessing the feasibility of Phase 3 vaccine trials against Marburg Virus Disease: a modelling study

**DOI:** 10.1101/2023.02.22.23286294

**Authors:** George Y Qian, Thibaut Jombart, W John Edmunds

**Affiliations:** Department of Engineering Mathematics, University of Bristol, United Kingdom; Centre for Mathematical Modelling of Infectious Diseases, London School of Hygiene & Tropical Medicine, London, United Kingdom; MRC Centre for Global Infectious Disease Analysis, Department of Infectious Disease Epidemiology, School of Public Health, Imperial College London, London, United Kingdom

## Abstract

**Background:** Outbreaks of Marburg virus disease (MVD) are rare and small in size, with only 16 recorded outbreaks since 1967, only two of which involved more than 100 cases. It has been proposed, therefore, that Phase 3 trials for MVD vaccines should be held open over multiple outbreaks until sufficient end points accrue to enable vaccine efficacy (VE) to be calculated. Here we estimate how many outbreaks might be needed for VE to be estimated.

**Methods:** We adapt a mathematical model of MVD transmission to simulate a Phase 3 individually randomised placebo controlled vaccine trial. We assume in the base case that vaccine efficacy is 70% and that 50% of individuals in affected areas are enrolled into the trial (1:1 randomisation). We further assume that the vaccine trial starts two weeks after public health interventions are put in place and that cases occurring within 10 days of vaccination are not included in VE calculations.

**Results:** The median size of simulated outbreaks was 2 cases. Only 0.3% of simulated outbreaks were predicted to have more than 100 MVD cases. 95% of simulated outbreaks terminated before cases accrued in the placebo and vaccine arms. Therefore the number of outbreaks required to estimate VE was large: after 100 outbreaks, the estimated VE was 69% but with considerable uncertainty (95% CIs: 0% - 100%) while the estimated VE after 200 outbreaks was 67% (95% CIs: 42% - 85%). Altering base-case assumptions made little difference to the findings.

**Conclusions:** It is unlikely that the efficacy of any candidate vaccine can be calculated before more MVD outbreaks have occurred than have been recorded to date. This is because MVD outbreaks tend to be small, public health interventions have been historically effective at reducing transmission, and vaccine trials are only likely to start after these interventions are already in place. Hence, it is expected that outbreaks will terminate before, or shortly after, cases start to accrue in the vaccine and placebo arms. Manufacturers may want to consider alternative routes to licensure than Phase 3 trials for MVD vaccines.

## Introduction

Marburg virus disease (MVD) is an acute, highly pathogenic, zoonotic, haemorrhagic disease caused by infection with Marburg virus. The wild reservoir of the Marburg virus is the Egyptian fruit bat (*Rousettus aegyptiacus*), which has a wide geographical range covering many parts of sub-Saharan Africa and the Middle East (1). The virus was first discovered in a laboratory-derived outbreak in Marburg, West Germany in 1967 (2). Since then, there have been 15 other spillover human infections or outbreaks of MVD, most of which have occurred in Sub-Saharan Africa, often associated with exposure to bats in caves or mines (3). Human-to-human transmission is possible and whilst most outbreaks have been small, two outbreaks (one in DRC in 1998-2000 and one in Angola in 2004-2005) resulted in hundreds of cases and many deaths (4,5).

The high pathogenicity associated with MVD and its potential to spread and cause public health emergencies has led the World Health Organization (WHO) to designate MVD as a priority for research and development into new vaccines, therapeutics and diagnostics (6). Accordingly, there is now a number of MVD vaccine candidates which are in pre-clinical testing and one (a Chimpanzee Adenovirus vectored vaccine produced by the Sabin Vaccine Institute) which has completed a Phase 1 trial in humans (7) with Phase 2 trials planned for 2023. It is widely acknowledged, however, that Phase 3 efficacy trials for MVD vaccines will be very challenging to conduct due to the sporadic nature of MVD outbreaks which may occur over a large geographical range, the relatively small size of most outbreaks, and the necessity to control them as rapidly as possible using existing public health and infection control measures. Any Phase 3 vaccine trial will therefore have to deploy quickly to an affected area and even then the epidemic may end before sufficient cases have accumulated to determine vaccine efficacy with a degree of statistical confidence. Given the scarcity of MVD outbreaks there may be pressure to evaluate a number of available vaccine candidates simultaneously, with the potential of further eroding statistical power if different trials competed for eligible participants during an outbreak. To help negate these problems, an endpoint-driven platform trial design has been proposed (8) under a master protocol approach which could potentially remain open for recruitment over multiple outbreaks until sufficient endpoints have accrued.

Nevertheless, questions remain: how many outbreaks might be needed to evaluate an MVD vaccine, and thus, given the low frequency of MVD outbreaks, how feasible are Phase 3 trials likely to be? This study aims to answer these questions by utilising a mathematical model of MVD, which was parameterised based on a systematic review of data from all 16 previous MVD outbreaks (3). The model is used to simulate an hypothetical individually randomised Phase 3 MVD vaccine trial to determine how many cases and outbreaks might be required to determine vaccine efficacy (VE).

## Methods

We used a branching process model previously developed to simulate MVD transmission over time. New infections generated at any time *t* are governed by the force of infection *λ*_*t*_, which is determined by previous case incidence *y*_*s*_ (*s* = 1, …, *t*-1), the serial interval distribution (denoted by *w*, its probability mass function), and the reproduction numbers *R*_*s*_ as:

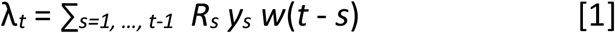

New secondary cases at time *t* are then drawn from a Poisson distribution so that:

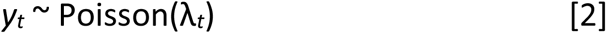

Equation [1] shows that the reproduction number *R*_*s*_ is allowed to vary over time. This is used to distinguish, in any given outbreak, three phases: a first one, during which transmission is maximum (*R*_*s*_ = *R*_0_, the basic reproduction number), a second one during which non-pharmaceutical intervention reduces transmission by a factor *E*, the intervention efficacy, so that:

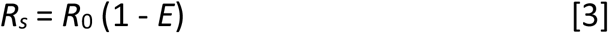

and finally, a third during which vaccination further reduces transmission by a factor *V*, the vaccine efficacy, so that:

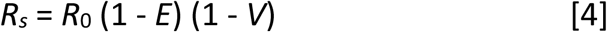

Intervention is defined, in this context, as the implementation of measures such as case isolation, contact tracing and barrier nursing. Vaccination is assumed to occur after intervention and so we modified the model to include a delay from intervention to vaccination.

Using this model, we simulated a trial involving a candidate vaccine with a nominal vaccine efficacy (VE) of 70% in the base case. We assumed a reactive mass vaccination strategy with 50% trial coverage in affected areas in the base case (9). Across all previous MVD outbreaks, the median delay between onset of the first case and beginning of interventions was 21 days (3). We assumed that 2 further weeks were required for a vaccination campaign to be implemented in the base case. Hence, interventions and mass vaccination were simulated 21 and 35 days, respectively, after the first case. The time for vaccine efficacy to peak was assumed to be 7 days [see (3)]. We assumed the rate of zoonotic introductions followed a beta distribution with parameters *α* = *0*.*01* and *β* = *4*, such that around 95% of the rate of introductions was under 1 per year, since most outbreaks had only one zoonotic case (see Appendix, Figure A1).

Trial participants were divided into 2 groups: one whose participants received the vaccine (25% of the population) and another who received a placebo (25% of the population). The other 50% of the population were assumed not to be enrolled in the trial, for various reasons *e*.*g*. ineligibility, refusal to consent, being absent on the day of enrolment etc. (9). We varied the fraction enrolled in the trial in the sensitivity analysis by simulating both 30% and 70% coverage. We used our branching process model with mass vaccination, described above and in (3) to simulate 5000 outbreaks of MVD. For each outbreak, we reported the total number of cases as well as the number of cases in the placebo and vaccine arms. Subsequently, we calculated the vaccination efficacies (10) and associated 95% confidence intervals for sets of 10, 20, 30 etc. outbreaks, using 500 bootstrap samples to account for the variation in outbreak sizes across simulations. Vaccine efficacy was defined as:

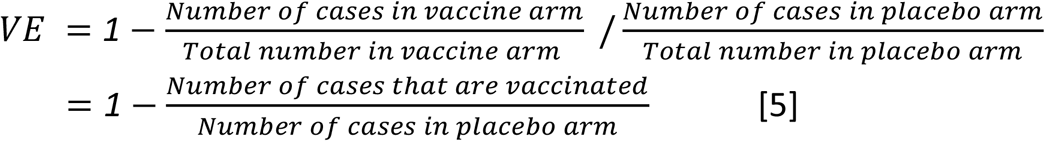

The second equality is due to having a balanced population in the placebo and vaccine arms.

Cases were included in the calculation of vaccine efficacy only if they occurred at least 10 days after vaccination, as it is assumed that there would be a delay between vaccination and the onset of immunity (9). Due to relatively low transmissibility of MVD and high efficacy of non-pharmaceutical interventions (NPIs) (3), some simulated outbreaks may be controlled before having any case in both the vaccine and placebo arms. These outbreaks were ignored for VE calculations, but our results reporting VE estimations by numbers of outbreaks were corrected for the proportion of outbreaks controlled solely with NPIs. To account for the uncertainty in some key parameters, we also carried out a sensitivity analysis, by changing the delay from the first MVD case to vaccination (from 35 days to 14 and 90 days), the vaccine coverage (from 50% to 30% and 70%), nominal efficacy (from 50% to 30% and 70%) and time from vaccination to infection in those that are infected (from a median of 9 days to 20 days). We also simulated a ‘best case’ scenario in which we decreased the delay from the first MVD case to vaccination to 14 days, increased the coverage to 70%, and increased the time from vaccination to infection to a median of 20 days.

## Results

Given a vaccine with a nominal 70% VE, and a vaccination campaign beginning 35 days after the first case was detected, we found that the median outbreak size was 2 cases, and the maximum 323. Only 0.3% of simulated outbreaks were predicted to have more than 100 MVD cases. For the vast majority (97%) of simulated outbreaks, there were no cases in the vaccine arm, *i*.*e*. the majority of outbreaks would be controlled in the absence of a vaccine. Furthermore, in 95% of all outbreaks, there were no cases in both the placebo and vaccine arms. The largest outbreak (323 cases) had 10 and 36 cases in its vaccine and placebo arms, respectively.

Figure 1 shows the estimated vaccine efficacies and their 95% CIs as a function of the number of outbreaks, including those where there were zero cases in both the vaccine and placebo arms. When VE was calculated using fewer than 50 outbreaks, the median VE ranged from 68-73%, although the associated confidence intervals were wide (95% CIs: -∞%-100%). Note that VE can be -∞ if there are no cases in the placebo group but at least one case in the vaccine group, given equal numbers in the two arms. After 100 outbreaks, the VE estimate was 69% (95% CIs: 0% - 100%) while the estimated efficacy after 200 outbreaks was 67% (95% CIs: 42% - 85%).

**Figure 1:**
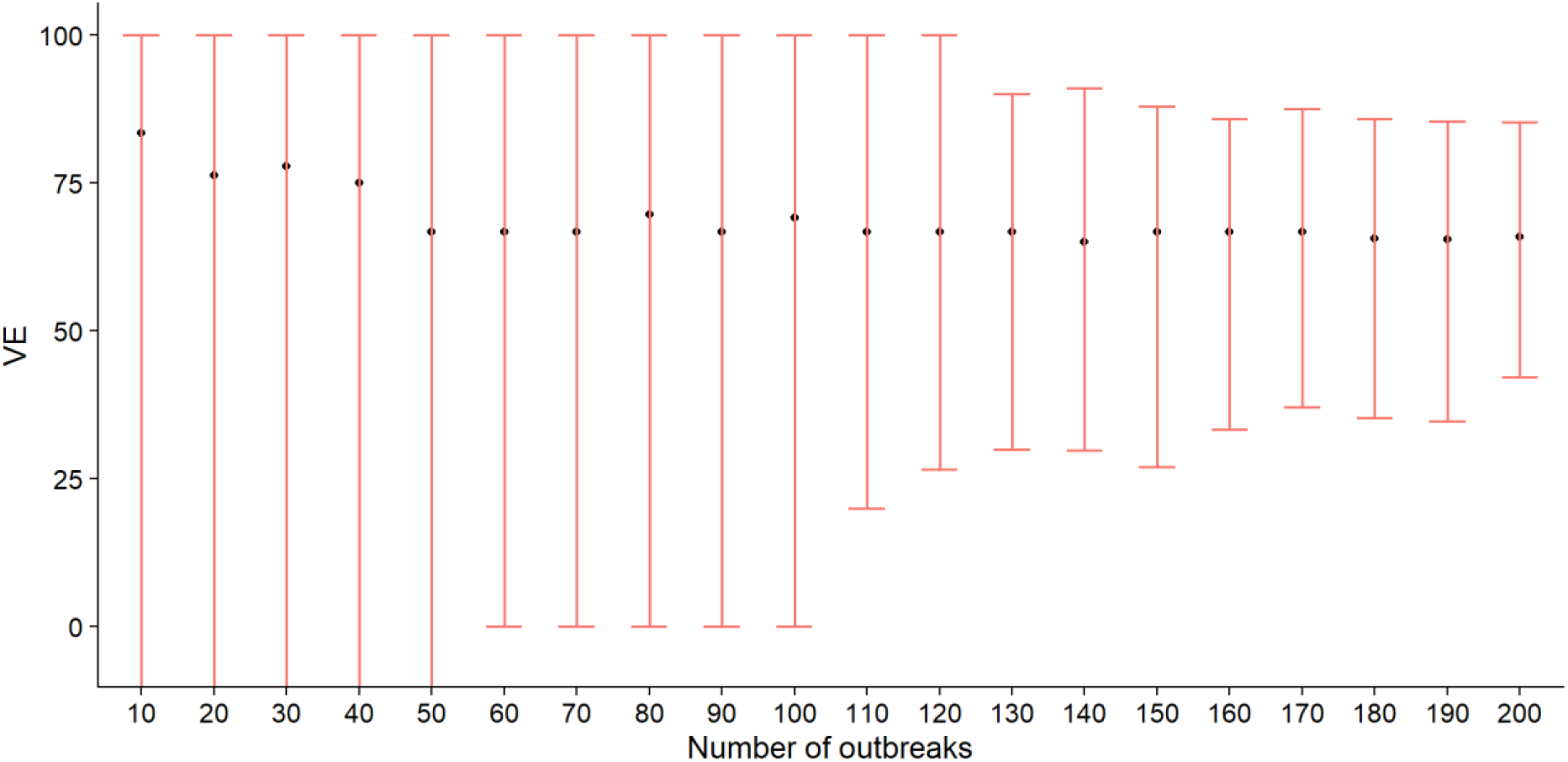
**Vaccine efficacies and associated 95% bootstrapped confidence intervals calculated after different numbers of outbreaks (including those where there were 0 cases in both the vaccine and placebo arms). The lower CIs of the first 50 outbreaks were estimated to be negative**.

Table 1 shows the variation in estimated VE from our sensitivity analyses (adjusting the delay to vaccination, coverage and nominal vaccine efficacy). Across all scenarios where we changed only one parameter, confidence intervals for VE were -∞-100% after 10 outbreaks and remained wide even after 100 outbreaks (typically 0-100%). In general, scenarios involving a higher coverage, lower VE, earlier intervention/vaccination and a lengthier time between vaccination and infection reduced the CIs slightly. These scenarios also took fewer outbreaks to reach 150 cases in the vaccinated and placebo arms (see Table 2). In particular, lengthening the delay between vaccination and infection reduced the number of outbreaks required to reach 150 cases to 387 (95% CIs: 156-696). Moreover, a combination of these scenarios reduced the CIs of the VE estimates (57-80% after 200 outbreaks), as well as the number of outbreaks required to reach 150 cases (264 outbreaks (95% CIs: 107-490)) even further.

**Table 1:**
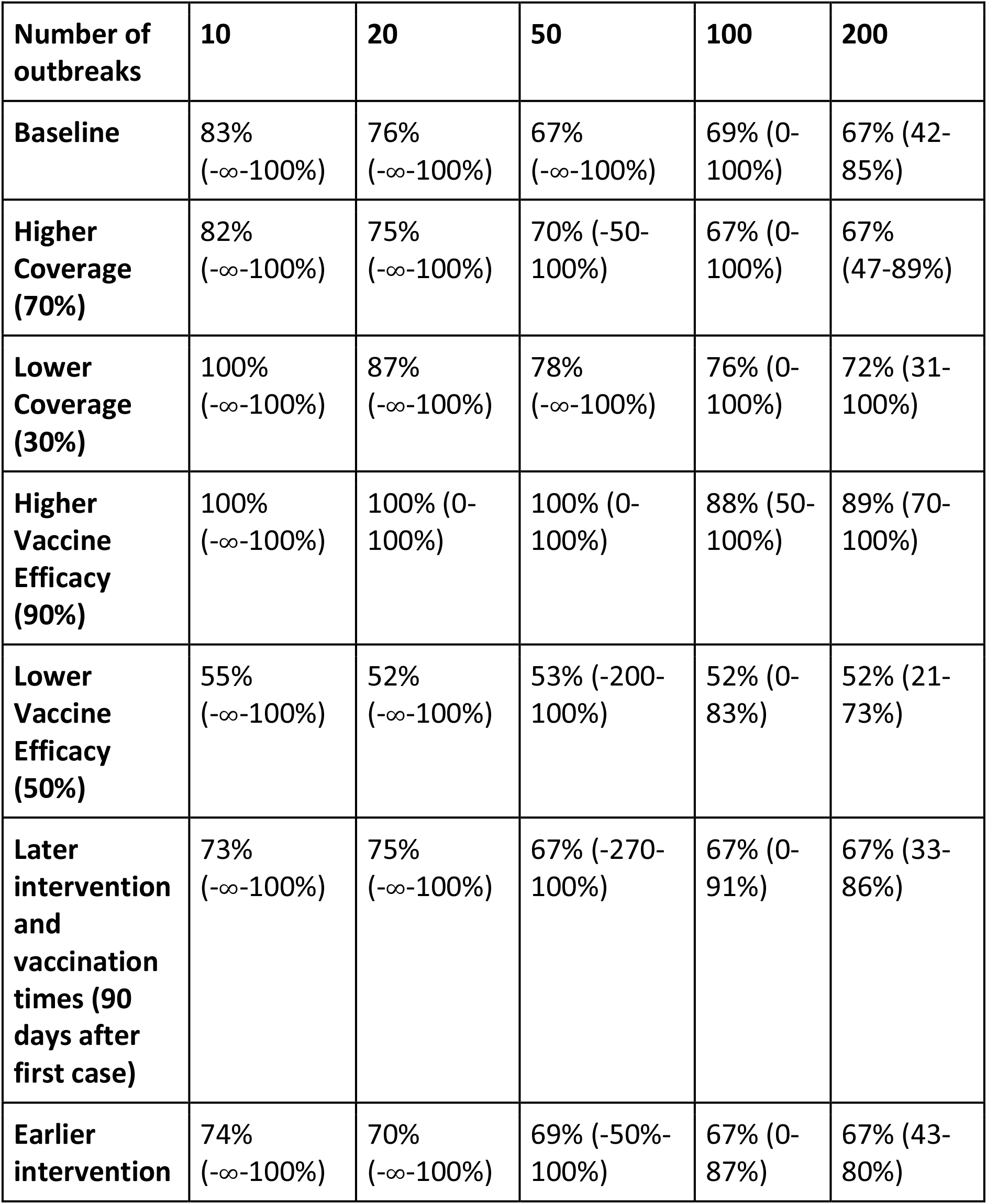
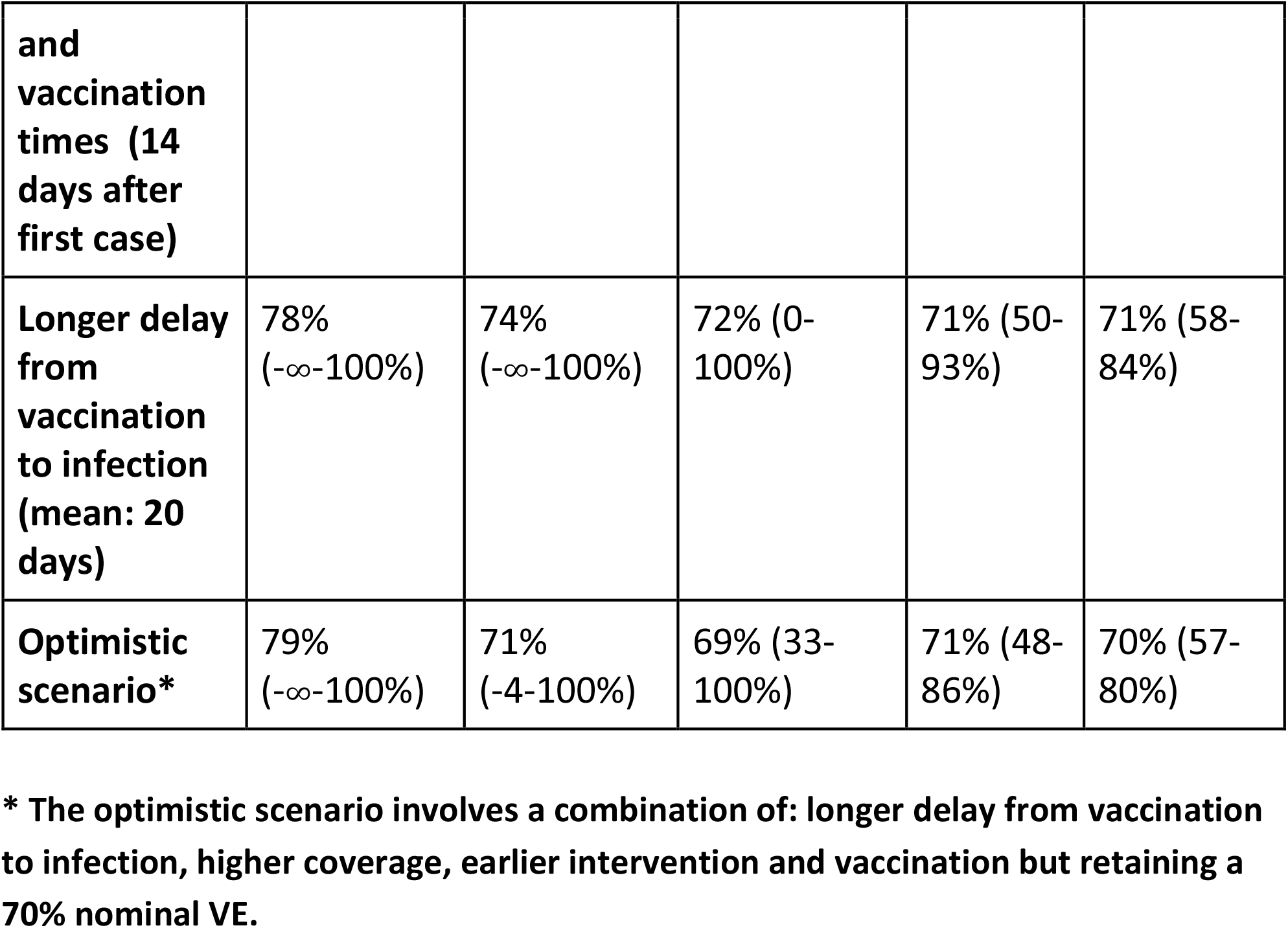
**Estimated median vaccine efficacy (VE) with 95% CIs (in brackets) calculated after 10, 20, 50, 100 and 200 simulated outbreaks, by varying the vaccination coverage, nominal VE and intervention/vaccination times. Baseline values: 50% coverage, 70% nominal VE, intervention and vaccination at 21 and 35 days, respectively**.

**Table 2:**
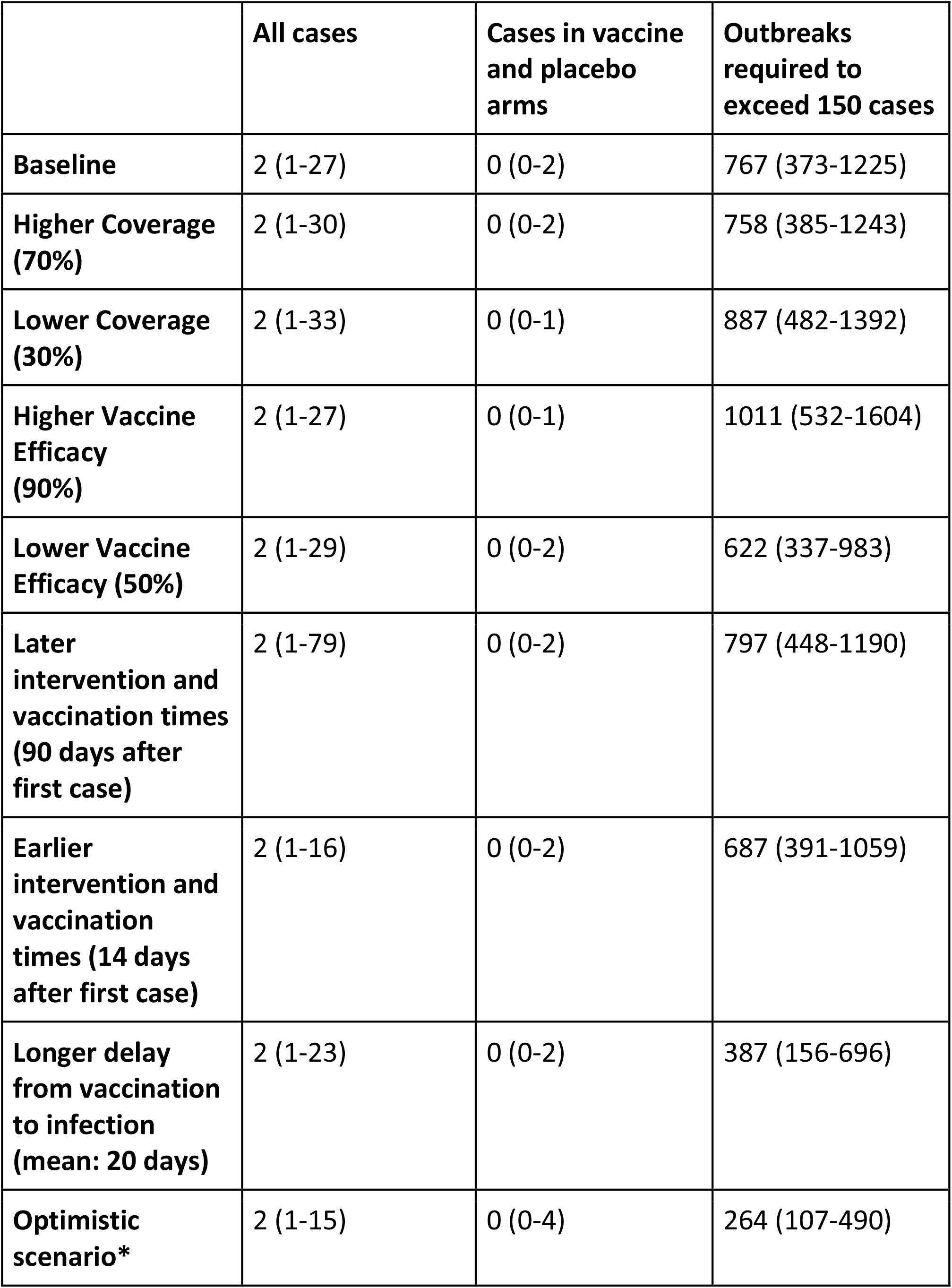
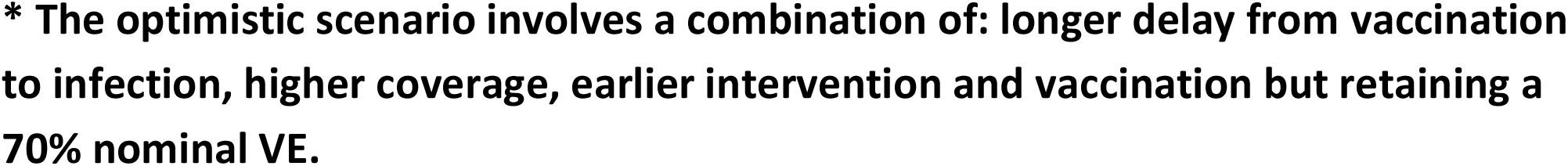
**Median number of MVD cases overall, median number of cases in both arms and average number of outbreaks required to exceed 150 cases in both arms, with 95% CIs (in brackets) under different scenarios**.

|  | **All cases** | **Cases in vaccine and placebo arms** | **Outbreaks required to exceed 150 cases** |
| --- | --- | --- | --- |
| **Baseline** | 2 (1-27) | 0 (0-2) | 767 (373-1225) |
| **Higher Coverage (70%)** | 2 (1-30) | 0 (0-2) | 758 (385-1243) |
| **Lower Coverage (30%)** | 2 (1-33) | 0 (0-1) | 887 (482-1392) |
| **Higher Vaccine Efficacy (90%)** | 2 (1-27) | 0 (0-1) | 1011 (532-1604) |
| **Lower Vaccine Efficacy (50%)** | 2 (1-29) | 0 (0-2) | 622 (337-983) |
| **Later intervention and vaccination times (90 days after first case)** | 2 (1-79) | 0 (0-2) | 797 (448-1190) |
| **Earlier intervention and vaccination times (14 days after first case)** | 2 (1-16) | 0 (0-2) | 687 (391-1059) |
| **Longer delay from vaccination to infection (mean: 20 days)** | 2 (1-23) | 0 (0-2) | 387 (156-696) |
| **Optimistic scenario\*** | 2 (1-15) | 0 (0-4) | 264 (107-490) |

**\* The optimistic scenario involves a combination of: longer delay from vaccination to infection, higher coverage, earlier intervention and vaccination but retaining a 70% nominal VE.**

## Discussion

It is unlikely that the efficacy of any candidate vaccine could be calculated before more MVD outbreaks have occurred than have been recorded to date. Our results suggest that the vast majority of future outbreaks may have no cases in both the vaccine and placebo arms, and would in fact be controlled before VE estimations could begin (ten days after implementing the vaccination campaign, which is when cases are included in any VE calculations). NPIs were sufficient to control 95% of simulated outbreaks. This is consistent with historical data: although MVD has a high case-fatality ratio, it is a rare and sporadic disease, with only 16 known outbreaks since 1967, and a low but variable reproduction number (varying across outbreaks from 0.5 [95% CI: 0.05 – 1.8] to 1.2 [95% CI: 1.0 – 1.9]), becoming substantially lower once NPIs have started (varying across outbreaks from 0.2 [95% CI: 0.006 – 0.7] to 0.6 [95% CI: 0.03 – 1.5]) (3). Additionally, any candidate vaccine itself would offer protection against MVD symptoms (70% VE in the base case), further limiting the size of any outbreak.

It was previously calculated that 150 cases, both in the vaccine and control arms, would be required to estimate VE (8). Figure 2 shows a histogram of the number of simulated MVD outbreaks required to reach 150 combined cases in the vaccine and placebo arms. The median number of outbreaks was 767 and even in the best-case scenario under baseline conditions, 186 outbreaks were required, which is an order of magnitude higher than the 16 MVD outbreaks recorded to date.

**Figure 2:**
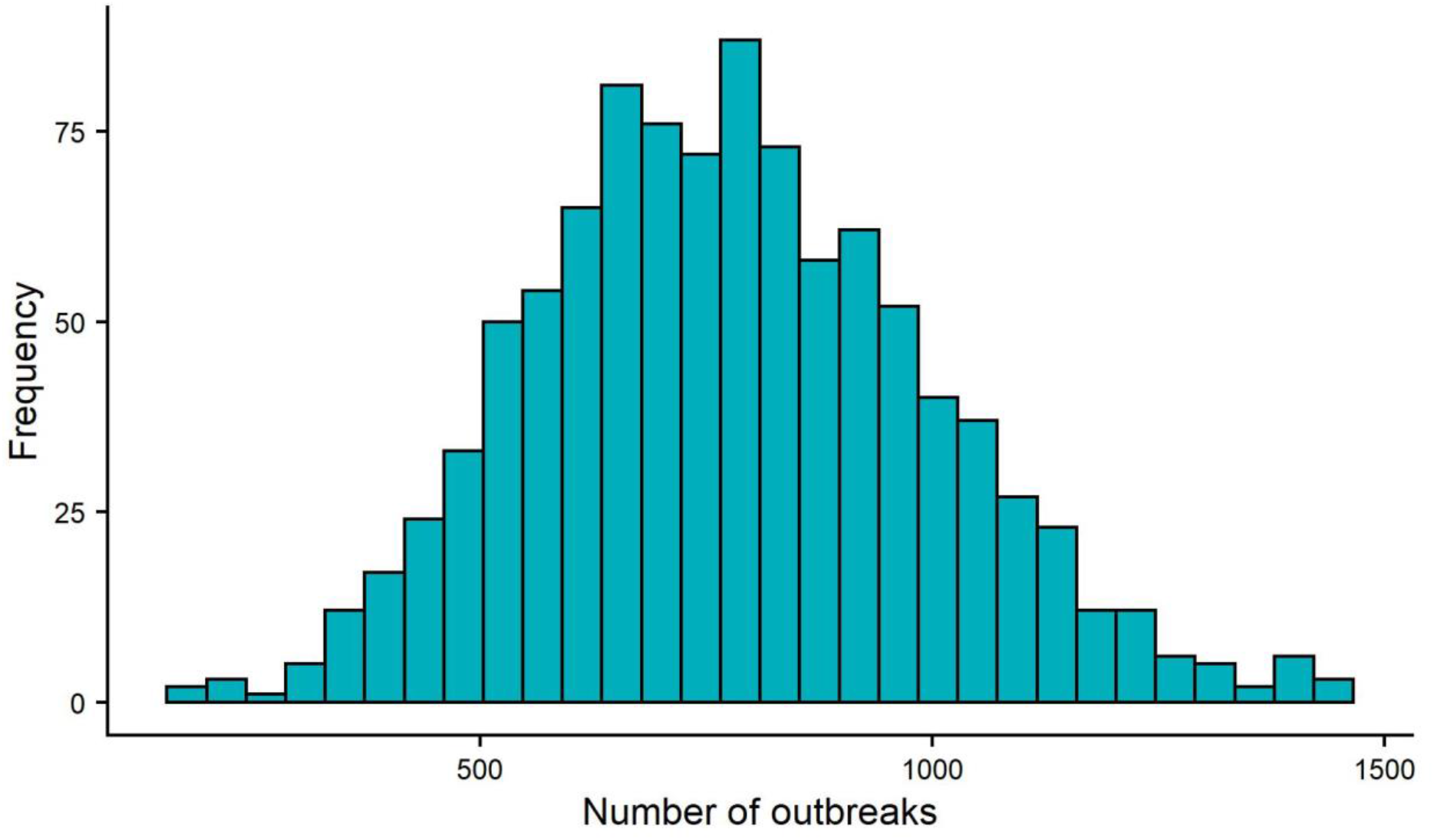
Number of simulated outbreaks required to reach 150 total cases for the vaccine and placebo arms, based on 1000 simulations.

Furthermore, while there were over 150 cases in two previous outbreaks (154 in DRC, 1999-2000 and 374 in Angola, 2004-5), a large proportion of these cases occurred before effective interventions had taken place (82 and 142 cases, respectively) (3). In a clinical trial, cases occurring before interventions would not be included in VE calculations, since the vaccination trial would not yet have been set up. Table A1 (Appendix) shows the number of cases from the 16 recorded MVD outbreaks, both overall and after interventions were put in place. 12 outbreaks had either 0 or 1 case after intervention, while 2 further outbreaks had 5 cases post-interventions. Only the 2 large outbreaks (DRC and Angola) had more than 5 cases post intervention, and these were likely due to repeated zoonotic transmission in DRC (4) or delayed adherence to the intervention programme in Angola (5). Finally, detection of MVD outbreaks and intervention efficacy have improved since these two outbreaks (11), suggesting that future outbreaks may be more quickly contained than in the past.

Our sensitivity analyses (see Tables 1 and 2) show that while varying parameters such as an increased coverage rate, VE, decreased time to intervention/vaccination and, in particular, lengthening the delay from vaccination to infection did reduce the number of outbreaks required to accurately determine VE, this number was still in the hundreds, on average. Even after 200 outbreaks, the difference between the upper and lower confidence intervals ranged from 20-70%, depending on the scenario simulated. These CIs reflect the very large heterogeneity in outbreak size distribution and reinforce our message that obtaining the VE of any candidate MVD vaccine will be difficult in a Phase 3 trial, even if using the master protocol approach (8).

There are several limitations to our study. First, no licensed vaccine yet exists and so parameters such as the nominal VE are indicative only. There is also a general paucity of data available on the epidemiology of MVD and the effectiveness of public health interventions designed to reduce its spread, due to the sporadic nature of MVD outbreaks. In addition, we assume that both interventions and vaccine trials were implemented across all affected areas immediately, whereas in reality they may be rolled out over time. Using time-varying functions might help improve the accuracy somewhat, but there is a lack of data to inform the modelling of these functions. While this would decrease the number of outbreaks required for estimating VE, our conclusion that a prohibitively large number of outbreaks would be required is almost certainly robust to such changes.

## Conclusions

Our simulations suggest that a Phase 3 vaccine trial run under the master protocol approach where endpoints are accumulated over multiple outbreaks, would likely require a large number of outbreaks to accurately estimate vaccine efficacy - more outbreaks than have been observed since MVD was first discovered in 1967. Manufacturers should consider alternative routes to licensure than Phase 3 trials for MVD vaccines.

## Data Availability

All data produced in the present study are available upon reasonable request to the authors

## Acknowledgements

This study was part funded by the Department of Health and Social Care using UK Aid funding and is managed by the National Institute for Health and Care Research (VEEPED: PR-OD-1017-20002) and the Japan Agency for Medical Research and Development (AMED; grant number JP223fa627004). TJ acknowledges funding from the MRC Centre for Global Infectious Disease Analysis (reference MR/R015600/1), jointly funded by the UK Medical Research Council (MRC) and the UK Foreign, Commonwealth & Development Office (FCDO), under the MRC/FCDO concordat agreement and is also part of the EDCTP2 programme supported by the European Union. The views expressed in this publication are those of the authors and not necessarily those of the funders.

## Appendix

**Figure A1:**
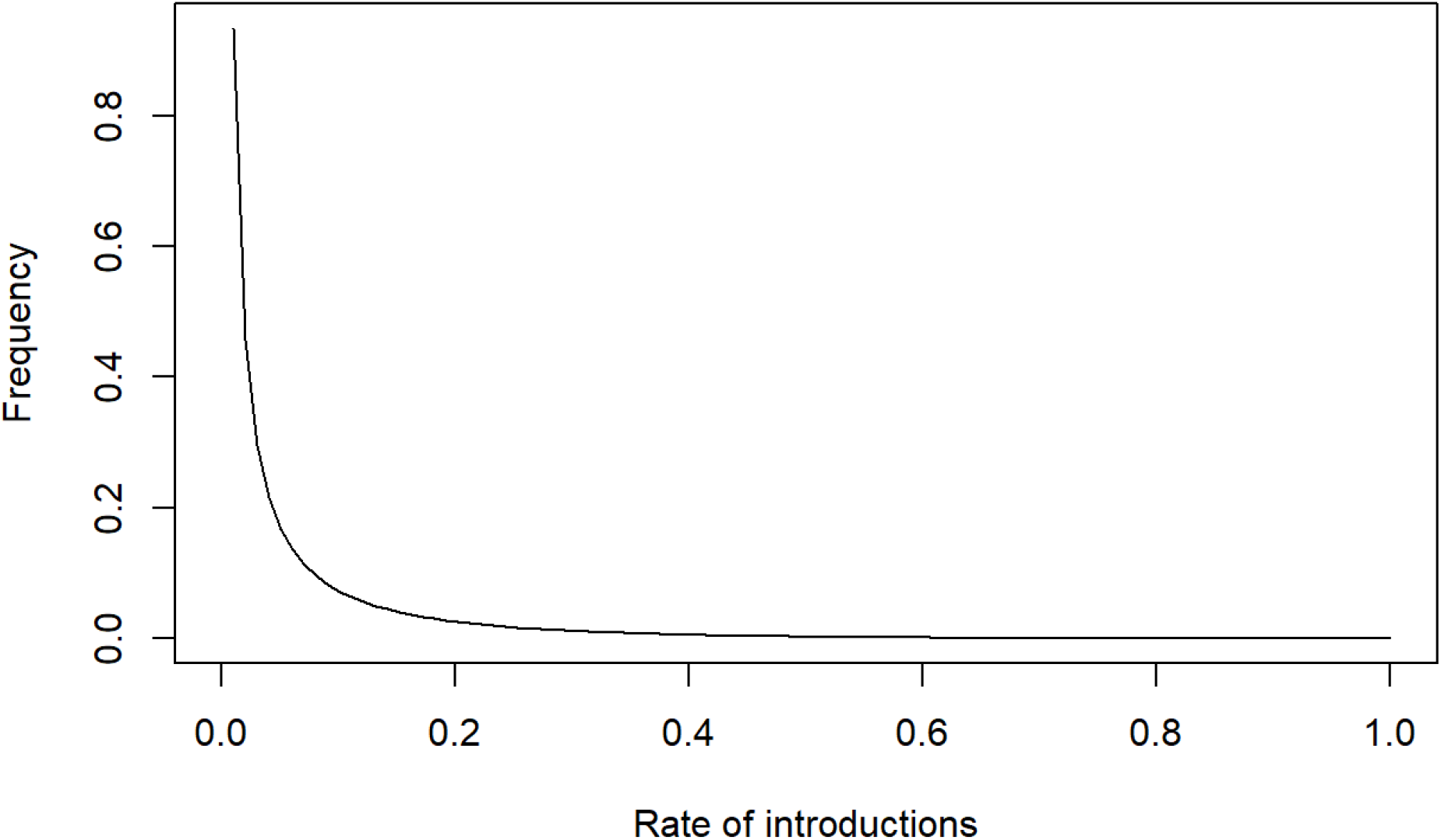
Plot showing the beta distribution of the rate of introductions per day used in our simulations.

**Table A1:** Marburgvirus outbreaks since 1967, including the number of cases both overall and after interventions were put in place.

| **Outbreak Location** | **Year(s)** | **Total number of cases** | **Cases after intervention** |
| --- | --- | --- | --- |
| **Marburg, Germany** | 1967 | 24 | 5 |
| **Frankfurt, Germany** | 1967 | 6 | 1 |
| **Belgrade, Yugoslavia** | 1967 | 2 | 1 |
| **Johannesburg, South Africa** | 1975 | 3 | 1 |
| **Nairobi, Kenya** | 1980 | 2 | 0 |
| **Nairobi, Kenya** | 1987 | 1 | 0 |
| **Durba and Watsa, Democratic Republic of Congo** | 1998-2000 | 154 | 72 |
| **Uige, Angola** | 2004-2005 | 374 | 232* |
| **Uganda** | 2007 | 4 | 1 |
| **USA (via Uganda)** | 2008 | 1 | 0 |
| **Netherlands (via Uganda)** | 2008 | 1 | 0 |
| **Uganda** | 2012 | 26 | 5 |
| **Uganda** | 2014 | 1 | 0 |
| **Uganda** | 2017 | 4 | 0 |
| **Guinea** | 2021 | 1 | 0 |
| **Ghana** | 2022 | 4 | 1 |

\* The interventions in Angola were reportedly slow to be adopted

